# Mammo-Bench: A Large-scale Benchmark Dataset of Mammography Images

**DOI:** 10.1101/2025.01.31.25321510

**Authors:** Gaurav Bhole, S Suba, Nita Parekh

**Author notes:** {, } and.

## Abstract

Breast cancer remains a significant global health concern, and machine learning algorithms and computer-aided detection systems have shown great promise in enhancing the accuracy and efficiency of mammography image analysis. However, there is a critical need for large, benchmark datasets for training deep learning models for breast cancer detection. In this work we developed Mammo-Bench, a large-scale benchmark dataset of mammography images, by collating data from six well-curated resources, *viz*., DDSM, INbreast, KAU-BCMD, CMMD, CDD-CESM and DMID. To ensure consistency across images from diverse sources while preserving clinically relevant features, a preprocessing pipeline that includes breast segmentation, pectoral muscle removal, and intelligent cropping is proposed. The dataset consists of 19,731 high-quality mammographic images from 6,500 patients across 6 countries and is one of the largest open-source mammography databases to the best of our knowledge. To show the efficacy of training on the large dataset, performance of ResNet101 architecture was evaluated on Mammo-Bench and the results compared by training independently on a few member datasets and an external dataset, VinDr-Mammo. An accuracy of 78.8% (with data augmentation of the minority classes) and 77.8% (without data augmentation) was achieved on the proposed benchmark dataset, compared to the other datasets for which accuracy varied from 25 – 69%. Noticeably, improved prediction of the minority classes is observed with the Mammo-Bench dataset. These results establish baseline performance and demonstrate Mammo-Bench’s utility as a comprehensive resource for developing and evaluating mammography analysis systems.

## 1 Introduction

Early detection of breast cancer through reliable screening methods such as mammography remains crucial for successful treatment outcomes, and hence mammogram screening programs have been established worldwide. The interpretation of mammograms, however, requires careful analysis of various abnormalities such as masses (dense regions with varying shapes or patterns), calcifications (calcium deposits), architectural distortions (irregular patterns in breast tissue structure) and asymmetries (differences between corresponding regions in paired mammograms). This complexity, combined with factors like radiologist fatigue and varying expertise levels across hospitals, can lead to incorrect diagnosis. Computer Aided-Detection (CAD) systems have emerged as valuable tools to assist radiologists in identifying and classifying suspicious lesions, potentially improving detection accuracy and reducing false positives. The effectiveness of CAD systems, particularly those based on machine learning algorithms, is fundamentally dependent on the quality and size of training data. Large number of high-quality mammography images are required to capture the diverse manifestations of abnormalities and provide comprehensive annotations that include abnormality types and clinical metadata such as breast density, BI-RADS scores and molecular subtype information. Balanced representation of different case types and diverse patient demographics are crucial for developing reliable diagnostic models. Recent studies [1] have shown that such comprehensive datasets are required for developing robust deep learning models that can effectively generalize across different clinical settings and patient populations.

While several public mammography datasets exist, they often have limitations that restrict their utility for developing comprehensive CAD systems. Common challenges include small numbers of samples, inconsistencies in image quality and annotations, and significant class imbalances. Additionally, most datasets are collected from single institutions, potentially limiting their generalizability across different populations and imaging protocols. A large benchmark mammography dataset is crucial in development of machine learning methods since it would provide a “baseline” for what current algorithms can achieve on a standardized set of images and offer a reference point for setting goals and measuring progress in the research applications.

After evaluating various factors such as size, image quality, annotation completeness, and clinical metadata of mammography datasets in the public domain, we selected six well-curated datasets: DDSM [2], INbreast [3], KAU-BCMD [4], CMMD [5], CDD-CESM [6] and DMID [7]. We constructed a large benchmark dataset, Mammo-Bench, that addresses the limitations of individual datasets by unifying and standardizing data from these six sources. Before merging the images from individual resources, a rigorous pre-processing pipeline is proposed to ensure consistency and improve quality of the dataset. Annotations such as BI-RADS scores, breast density, abnormalities and asymmetries, and molecular subtypes were extracted from the respective member databases (if available), and masks for Regions of Interest (ROI) were generated by us for all the images. Mammo-Bench is a collection of 19,731 high-quality mammographic images from 6,500 patients across 6 countries. To the best of our knowledge, there is only one another resource, IRMA*, with 10,509 images, which has collated data from four resources, *viz*., DDSM [2, 8], MIAS [9], LLNL* [10] and RWTH* [11]; however, it is not an open-source dataset.

In this work, we have constructed a large-scale unified mammography dataset by collating and standardizing data from six well-curated resources. To achieve this, we proposed a comprehensive preprocessing pipeline that incorporates uniform image background and data format, breast segmentation and provide masks for regions of interest (RoIs), pectoral muscle removal, and intelligent cropping to ensure image consistency. Further, annotations such as BI-RADS scores, breast density, abnormality types, and molecular subtypes (extracted from individual sources) are provided to aid in breast cancer diagnosis. The detailed summary of the attributes of publicly available datasets is given in Table 1.

**Table 1.**
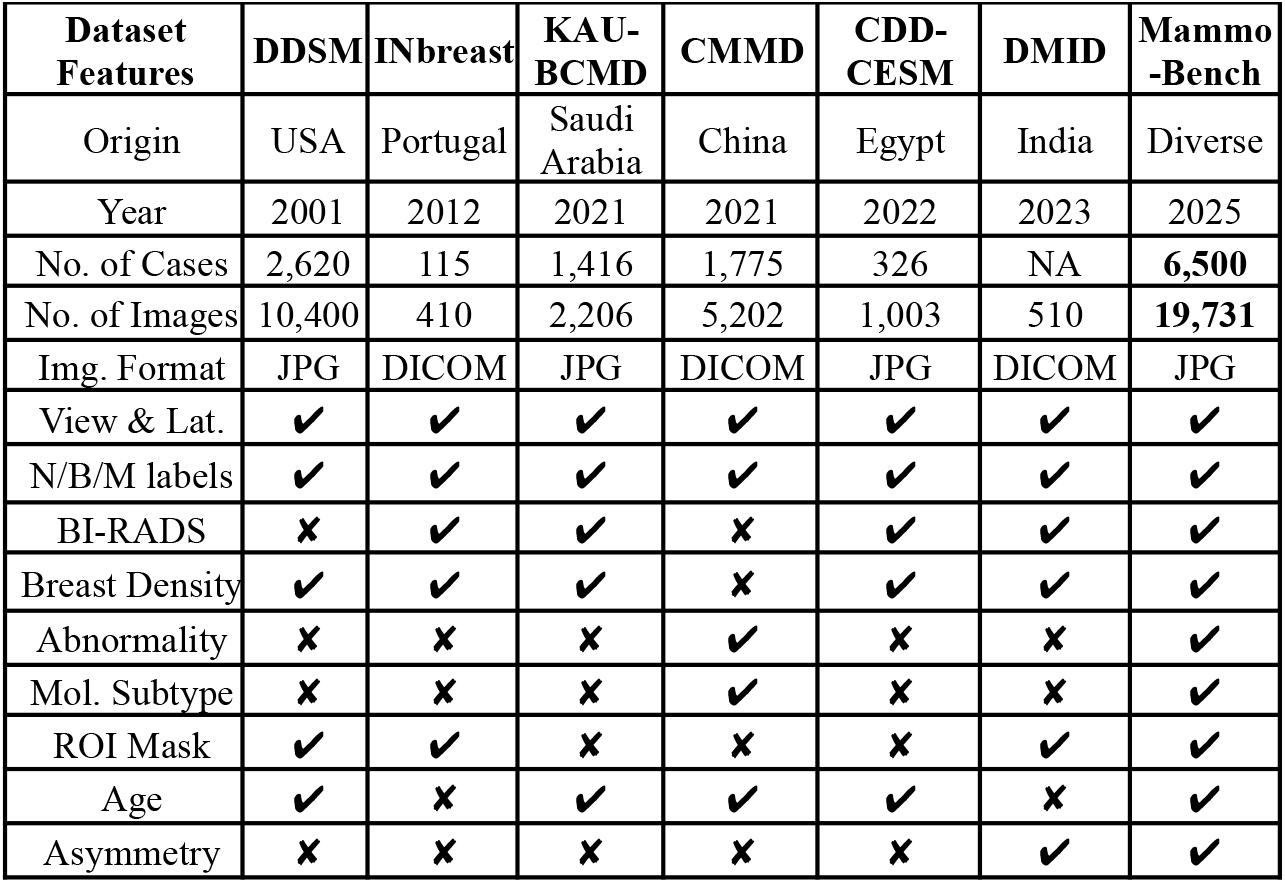
Summary of annotations and attributes of the six publicly available datasets considered for the construction of Mammo-Bench.

| Dataset Features | DDSM | INbreast | KAU-BCMD | CMMD | CDD-CESM | DMID | Mammo-Bench |
| --- | --- | --- | --- | --- | --- | --- | --- |
| Origin | USA | Portugal | Saudi Arabia | China | Egypt | India | Diverse |
| Year | 2001 | 2012 | 2021 | 2021 | 2022 | 2023 | 2025 |
| No. of Cases | 2,620 | 115 | 1,416 | 1,775 | 326 | NA | **6,500** |
| No. of Images | 10,400 | 410 | 2,206 | 5,202 | 1,003 | 510 | **19,731** |
| Img. Format | JPG | DICOM | JPG | DICOM | JPG | DICOM | JPG |
| View & Lat. | ✓ | ✓ | ✓ | ✓ | ✓ | ✓ | ✓ |
| N/B/M labels | ✓ | ✓ | ✓ | ✓ | ✓ | ✓ | ✓ |
| BI-RADS | ✗ | ✓ | ✓ | ✗ | ✓ | ✓ | ✓ |
| Breast Density | ✓ | ✓ | ✓ | ✗ | ✓ | ✓ | ✓ |
| Abnormality | ✗ | ✗ | ✗ | ✓ | ✗ | ✗ | ✓ |
| Mol. Subtype | ✗ | ✗ | ✗ | ✓ | ✗ | ✗ | ✓ |
| ROI Mask | ✓ | ✓ | ✗ | ✗ | ✗ | ✓ | ✓ |
| Age | ✓ | ✗ | ✓ | ✓ | ✓ | ✗ | ✓ |
| Asymmetry | ✗ | ✗ | ✗ | ✗ | ✗ | ✓ | ✓ |

To show the utility of the dataset, we performed various experiments. Three-class classification tasks (Normal, Benign, Malignant) with and without data augmentation, and a hierarchical binary classification strategy was implemented to address data imbalance. Our results demonstrate that this methodology significantly outperforms multi-class classification, suggesting a more robust path forward for CAD systems.

## 2 Related Works

Mammography datasets are vital resources for advancing breast cancer diagnosis and CAD systems and are categorized as open-source (e.g., DDSM, INbreast, etc.), or with restricted-access (e.g., OPTIMAM* [12], VinDr-Mammo* [13], etc.), depending on accessibility. Though well-curated, the publicly available mammographic datasets vary considerably in their coverage, size, annotations, quality, and accessibility, which affects their utility in diverse research applications. For example, image quality varies across datasets, for e.g., DDSM, MIAS and BancoWeb database* [14] contain older film-based digitized images, while recent datasets like CDD-CESM, LAMIS-DMDB* [15] provide high-quality digital mammograms. The size of the datasets is another critical issue, with INbreast, DMID and MIRacle dataset* [16] containing fewer images (< 500), while screening datasets like RSNA [17] contain thousands of images. Further, annotation quality and completeness vary widely across datasets, with ROI annotations available only for few datasets and that too not for the complete set (e.g., DDSM and DMID), impeding their use in lesion detection and localization tasks. Attributes such as breast density, BI-RADS scores, abnormality type, and molecular subtype are crucial in the accurate detection of breast malignancies. However, large variation is observed in the annotation details across the mammography datasets, for example, INbreast, KAU-BCMD, CDD-CESM, RSNA and DMID provide BI-RADS scores and breast density but no information about other abnormalities, while CMMD, and DMID provide abnormality type and molecular subtype annotations. Another factor of concern is data imbalance. The screening datasets such as RSNA, DDSM, NYU Screening Dataset* [18] and Cohort of Screen-Aged Women (CSAW)* [19] are large but highly imbalanced with the number of normal images (∼12×) compared to other classes. Most datasets include only mammography data while some provide multi-modal imaging data such as ultrasound, Tomosynthesis and MRI (e.g., OPTIMAM*) along with mammograms. Datasets also differ in terms of image formats, for example, RWTH dataset* (DICOM), BancoWeb* (TIFF), LLNL* (image cytometry standard (ICS)) and KAU-BCMD (JPG). Majority of mammography datasets are population specific with data collected from a single or multiple hospitals from the same demography, thereby limiting their use for developing generalizable models. Therefore, an open-access database consisting of large samples with complete annotations would be valuable for researchers developing AI models in breast cancer detection using mammograms.

## 3 Dataset Construction

### Description of the Dataset

The workflow for construction of Mammo-Bench, is given in Figure 1. First, data was downloaded from six mammography datasets: DDSM, INbreast, KAU-BCMD, CMMD, CDD-CESM and DMID (links provided in Supplementary Table 1). A brief description of the member databases is given below.

**Figure 1.**
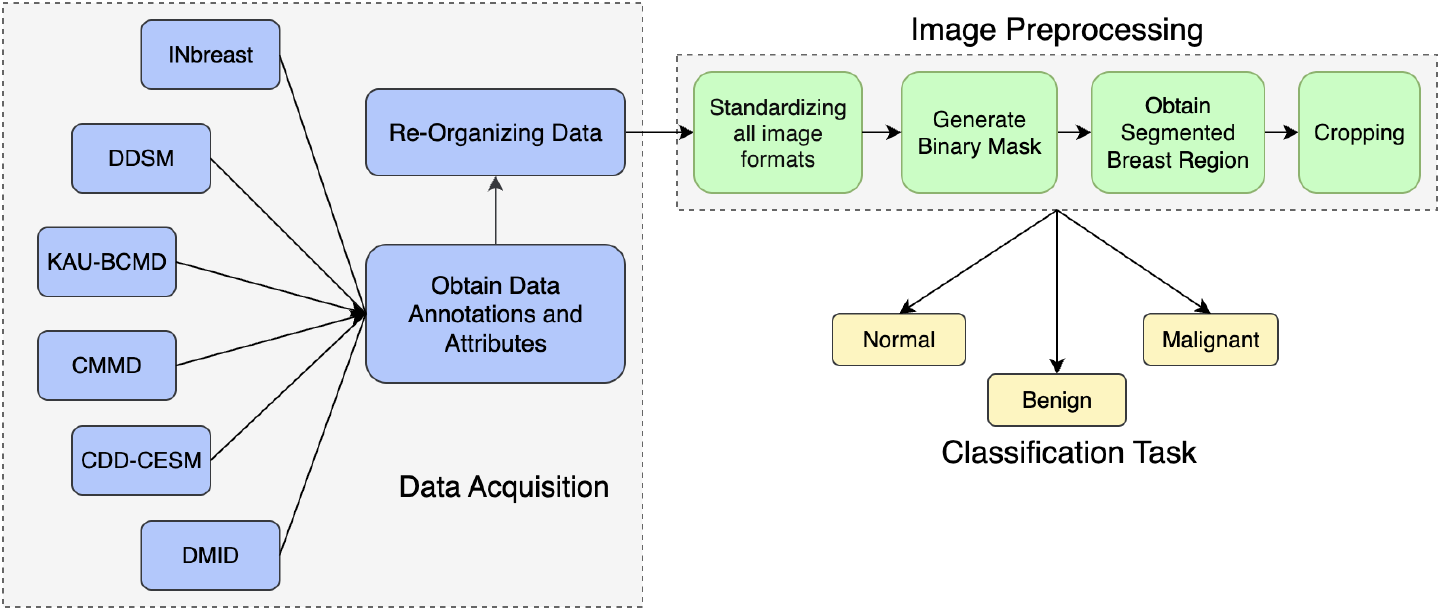
Overview of the Data Flow Pipeline

**DDSM**, the Digital Database for Screening Mammography [3] consists of 10,400 images from 2,620 patients in GIF format collected at Massachusetts General Hospital and Wake Forest University School of Medicine between 1990 - 1999. It offers a balanced distribution of 2,776 normal, 3,968 benign, and 3,656 malignant cases. Metadata provided includes cancer status (benign, malignant, or normal), patient age, and ROI boundaries for 2,830 images.

**INbreast dataset** [2], created by the breast research group at Centro Hospitalar de São João, Portugal, includes 115 cases with 410 digital mammogram images in DICOM format. This dataset provides detailed metadata, including BI-RADS scores, breast density, pixel-level lesion contours validated by two experts, and radiology reports. However, its major limitation is dataset size and class imbalance, with 67 normal, 243 benign, 43 suspicious malignant, and 57 malignant cases.

**KAU-BCMD**, the King Abdulaziz University Breast Cancer Mammogram Dataset [4] consists of 1,416 cases and 2,206 images in JPG format, collected between Apr’ 2019 – Mar’ 2020 at the Sheikh Mohammed Hussein Al-Amoudi Center of Excellence in Breast Cancer. Metadata includes patient age, previous screenings, breast density (manually assessed by radiologists), and BI-RADS scores (determined by three radiologists’ majority vote). About 172 images lack any annotation and were not included in the Mammo-Bench construction.

**CMMD**, the Chinese Mammography Database [5] comprises 5,202 images in DICOM format from 1,775 patients who underwent mammography examinations between July 2012 – Jan’ 2016, and is categorized into two subsets: CMMD1 (1,026 cases, biopsy-confirmed benign or malignant tumors) and CMMD2 (2,956 images with molecular subtype information: Luminal A, Luminal B, HER2-positive, and Triple-negative). This dataset includes age, benign/malignant labels, and abnormality type (mass, calcification or both), with images curated by two radiologists. Notably, it lacks ROI annotations but provides valuable molecular subtype data.

**CDD-CESM**, the Categorized Digital Database for Low Energy and Subtracted Contrast Enhanced Spectral Mammography [6] contains 1,003 low-energy images in JPG format and corresponding subtracted images, from 326 female patients collected between Jan’2019 – Feb’2021 at National Cancer Institute, Cairo University, Egypt. It is one of the balanced datasets with 341 normal, 331 benign, and 331 malignant cases.

**DMID**, the Digital Mammography Dataset for Breast Cancer Diagnosis Research [8], from Samved Hospital, India, contains 510 images in both DICOM and TIFF formats. It includes metadata on case types (normal, benign, malignant), BI-RADS scores, breast density, and abnormality type (calcification, mass and both). Each abnormality region is annotated with center coordinates, radius, and ROI mask. This dataset also provides balanced representation of normal and abnormal cases.

### Data Acquisition

After preliminary filtering of images with no available annotation (172 from KAU-BCMD), 19,731 images from ∼6,500 cases were considered for the construction of Mammo-Bench. Typically, four images per subject corresponding to two views for each breast are present: Mediolateral Oblique (MLO) and Craniocaudal (CC). The mapping between BI-RADS score and disease status labels is performed as given in Table 2 (according to [2, 4, 6, 8]) and all the cases with BI-RADS score 4a, 4b and 4c, have been relabeled to score 4. A summary of the annotations provided in the dataset is given in Table 2 and is briefly described below.

**Table 2.**
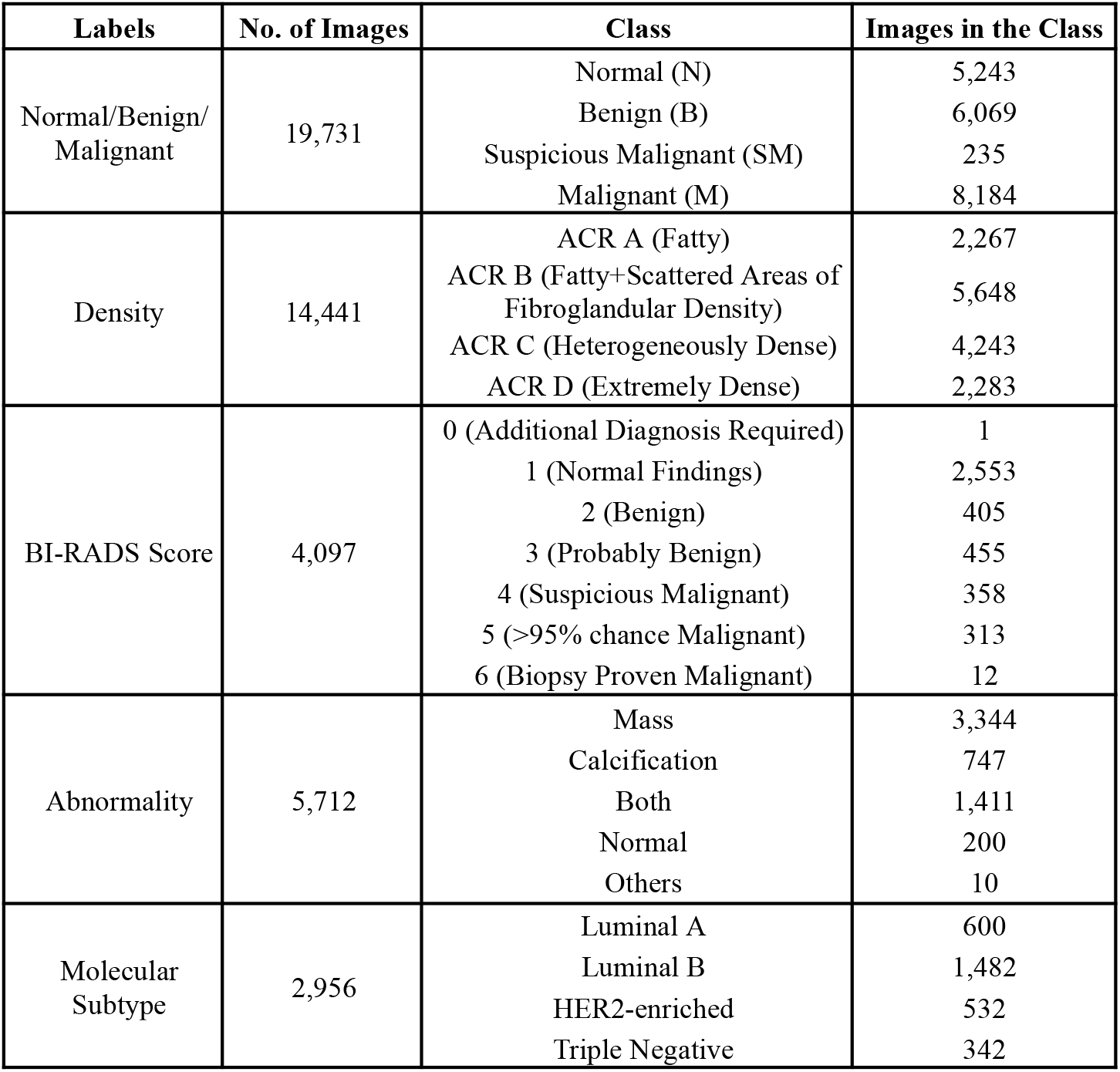
Description of various attributes of the images in the Mammo-Bench dataset.

#### Class Labels

Defines the disease status of the cases (images) as Normal, Benign, Suspicious Malignant, or Malignant.

#### Breast density

Breast density gives information about the different types of breast tissue, such as fat, fibrous, and glandular tissues. It is categorized as per the ACR standards into four categories A to D, as defined in Table 2.

#### BI-RADS Score

It is a standardized rating system used by radiologists to describe breast imaging test results and ranges from 0 to 6.

#### Abnormality

An abnormality is anything unusual found in breast tissue such as mass lumps or tiny calcium deposits.

#### Molecular Subtype

Based on the expression of the receptors, Progesterone (PR), Estrogen (ER), and human epidermal growth factor receptor 2 (HER2), breast cancer is grouped into four molecular subtypes, *viz*., Luminal A, Luminal B, HER2-positive, and Triple-negative, following the St. Gallen International Expert Consensus [20].

After the BI-RADS score – status label mapping, the number of images with Normal, Benign, Suspicious Malignant and Malignant class labels are 19,731 images (originally only 4,097 images had BI-RADS score). The number of images with breast density information is 14,441, all of which also have Normal (5,240), Benign (4,907), Malignant (4,061) and Suspicious Malignant (233) labels. Breast density annotation is valuable for studying relationships between breast density and cancer risk. Assessment of breast density is an important factor to the radiologists, as it is also used as a clinical indicator in deciding BI-RADS scores. It also helps in making informed decisions about additional screenings as dense breasts may occlude the presence of masses and calcified lesions in the mammography images making early detection of breast cancer challenging. Additionally, 5,712 images are annotated for specific abnormality types (mass, calcification, both), enabling detailed radiological assessment tasks. Molecular subtype information is available for only 2956 images from a single member database, CMMD. It helps mapping the morphological differences between the subtypes thereby providing a promising step towards precision medicine.

### Preprocessing

To improve the quality of mammography images collected from diverse datasets and ensure uniformity, following preprocessing steps were performed (Figure 2).

**Figure 2.**
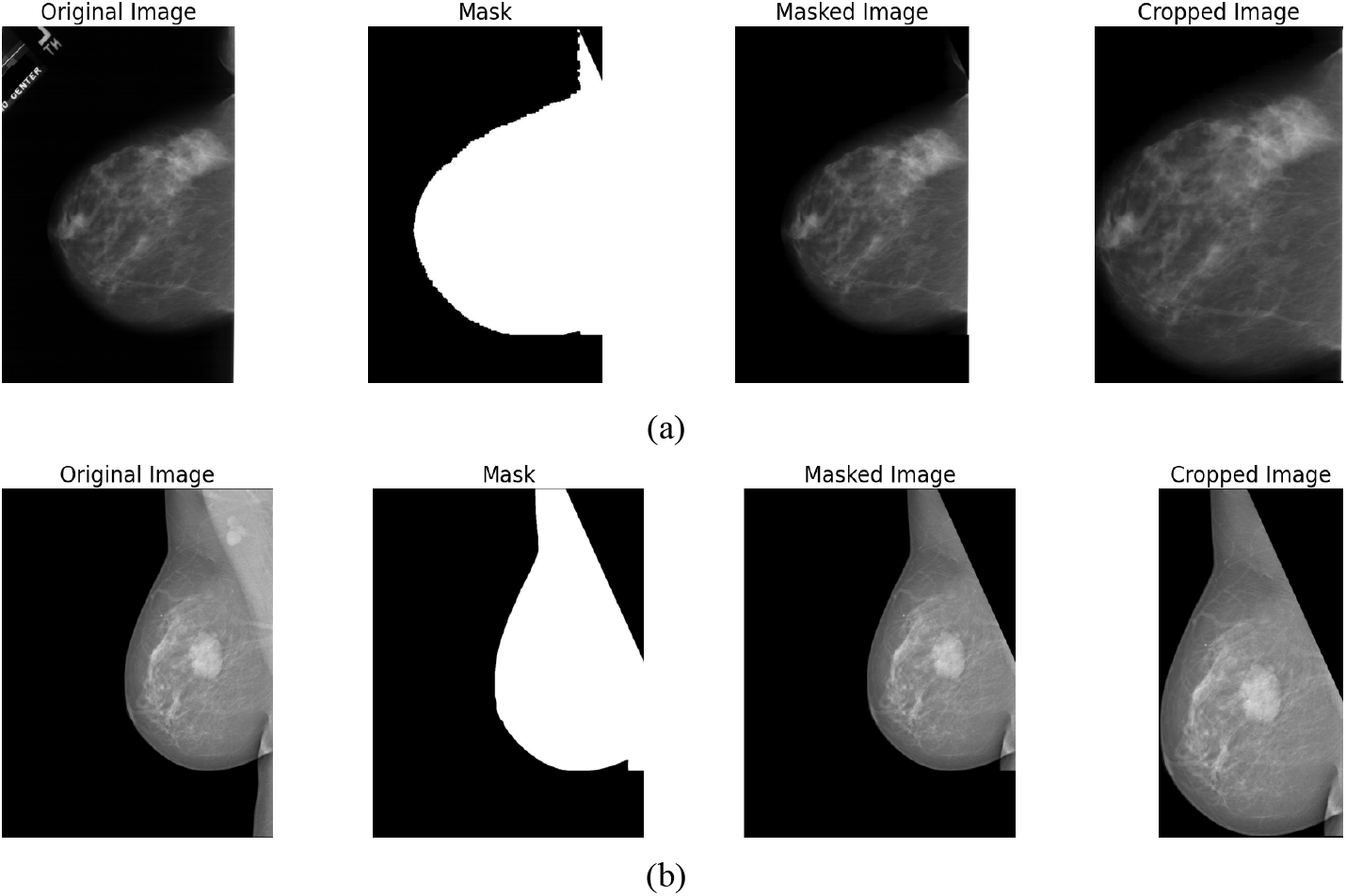
Illustration of the preprocessing pipeline: From left to right: Original mammogram, binary mask, breast region segmentation, and final cropped image for (a) Left-CC view (DDSM) and (b) Right-MLO view (INbreast). The effective removal of pectoral muscle in the MLO view and preservation of essential breast tissue in both views may be noted.

#### Data Format

Initially, all images were converted to JPG format from their original formats, and images with white backgrounds were standardized to black.

#### Breast Segmentation

The OpenBreast toolkit [21] was used to isolate the breast region through a multi-step segmentation process, as depicted in the second column of Figure 2. This process begins with background detection using a threshold-based method on the intensity histogram, followed by chest wall detection using a Hough-based line detector that represents edge pixels in parametric space. The process also ensures retention of the nipple region, identified as the farthest contour point from the detected chest wall.

#### Pectoral Muscle Removal

The pectoral muscle, commonly observed in MLO views and other irrelevant anatomical structures, which may confound in image analysis, are removed using the OpenBreast toolkit.

#### Mask Application

The segmentation process results in a binary mask for each image (third column in Figure 2). These masks effectively isolate the breast region while excluding irrelevant anatomical structures, and adapt to different breast shapes and orientations, both for CC views (Figure 2 (a)) and MLO views (Figure 2 (b)).

#### Image Cropping

An adaptive cropping algorithm based on contour detection [22] which removes extraneous background, retaining only the breast region is implemented (rightmost column in Figure 2). This algorithm applies a binary threshold determined through histogram analysis and empirical testing to identify the largest contour as the breast region, and crops the image along the computed bounding rectangle with appropriate padding to preserve the entire breast tissue.

### Organization of the Dataset

The data is organized into four main folders: ‘Original Dataset’ - contains the raw images, ‘Masks’ - generated breast region segmentation masks, ‘Preprocessed Dataset’ - processed images after applying the workflow (given in Fig 1), and ‘CSV Files’ - provide the clinical metadata. Each directory of images is organized into subfolders based on individual datasets. Within these subfolders, each image is assigned a unique identifier in the format ‘dataset_imgID.jpg’. A CSV file containing associated annotations from the member databases and paths to map images between Mammo-Bench and the individual datasets is provided. This is accompanied by helper codes in python for easy accessibility of the data. To support classification tasks with varied annotation labels, this file has been split into five separate CSV files, each tailored to specific classification tasks such as BI-RADS scores (0-6), breast density (A-D), abnormality )mass, calcification, both), and molecular subtype (Luminal A and B, Her2-enriched, Triple Negative). This organization of Mammo-Bench allows assessment of the impact of these attributes in classifying Normal, Benign, and Malignant cases.

## 4 Performance Evaluation

To assess the utility of the proposed dataset, Mammo-Bench, we performed the classification of the images (Normal, Benign, Malignant) using ResNet101 architecture pre-trained on the ImageNet database. Comparative performance evaluation was conducted by training ResNet101 on Mammo-Bench and three member datasets, *viz*., INbreast, CDD-CESM, DMID and one external dataset, VinDr-Mammo. First, to assess the impact of data imbalance, three experiments were performed using Mammo-Bench: three-class classification with and without data augmentation and hierarchical binary classification. Two data augmentation strategies were implemented on the minority class, namely, random rotation up to 10 degrees, and color jittering with brightness and contrast adjustments. Both transformations included resizing images to 224×224 and normalization using standard ImageNet statistics. For all experiments, the data was split into 80:20 for train-test sets, and performance was assessed using the metrics, precision, recall, F1-score, accuracy and Mathew’s correlation coefficient (MCC). In all cases the models were trained for 50 epochs, and the best weights were used for testing. The loss function used was categorical/binary cross-entropy and the optimizers were Adam and SGD.

### Three-Class Classification

In Table 3, performance of the ResNet101 model on Mammo-Bench dataset, with and without data augmentation, is given. As is seen from Table 3, there is only a marginal improvement in accuracy from 0.778 to 0.789 and MCC value from 0.59 to 0.61, on data augmentation. No difference in the precision (∼0.86) and recall (∼0.92) values of the normal class is observed, while marginal improvement in recall (0.74 to 0.77) for the malignant class and in precision (0.50 to 0.54) for the benign class is observed. This is probably due to the significant number of images in the two minority classes and ability of the deep architecture of ResNet101 in handling data imbalances to certain extent. For comparison, ResNet101 was independently trained on four datasets, *viz*., INbreast, CDD-CESM, DMID, and VinDr-Mammo and the averaged value of the performance metrics is given in Table 4. It may be noted that there is a significant drop in the performance of ResNet101 on these datasets, with accuracies ranging from 0.25 (DMID) to 0.69 (VinDr-Mammo). Though performance on VinDr-Mammo is reasonably good, it may be noted from Supplementary Table 1, that the recall of Benign and Malignant classes is ∼0, i.e., all the images are predicted as normal. These results clearly indicate the importance of a large benchmark dataset with good representation of all the classes for robust model building.

**Table 3.** Performance comparison of ResNet101 model for three-class classification without augmentation, with augmentation of minority classes and hierarchical binary classification using Mammo-Bench.

| Approach | Class | Precision | Recall | F1-Score | Accuracy | MCC |
| --- | --- | --- | --- | --- | --- | --- |
| Three-Class Classification | Normal | 0.865 | 0.928 | 0.895 | 0.778 | 0.595 |
|  | Benign | 0.502 | 0.369 | 0.425 |  |  |
|  | Malignant | 0.708 | 0.743 | 0.725 |  |  |
| Three-Class Classification with Augmentation of Minority Classes | Normal | 0.869 | 0.929 | 0.898 | 0.788 | 0.614 |
|  | Benign | 0.546 | 0.382 | 0.45 |  |  |
|  | Malignant | **0.709** | 0.777 | 0.741 |  |  |
| Hierarchical Binary Classification | Normal | **0.876** | **0.954** | **0.913** | 0.891 | 0.774 |
|  | Abnormal | 0.92 | 0.798 | 0.854 |  |  |
|  | Benign | **0.78** | **0.67** | **0.72** | 0.736 | 0.479 |
|  | Malignant | 0.7 | **0.81** | **0.75** |  |  |

**Table 4.**
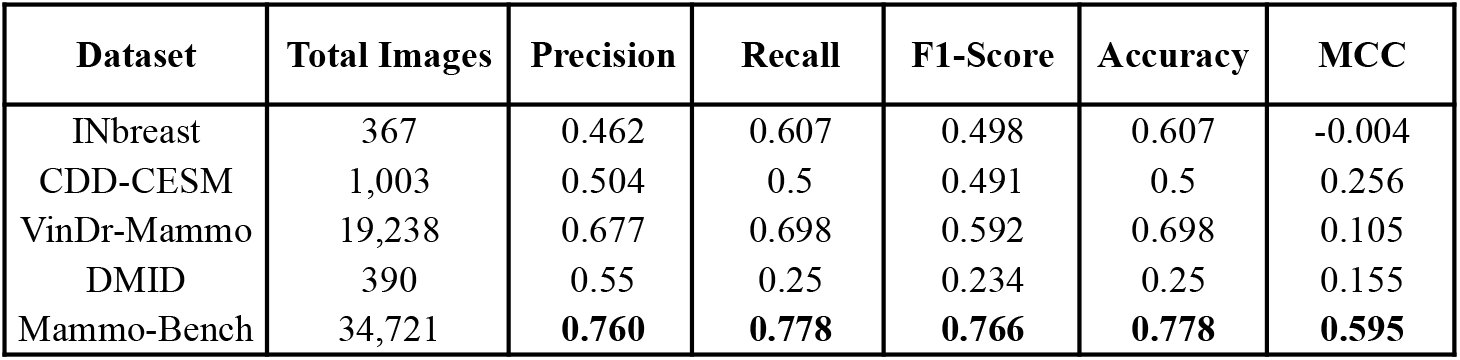
Performance comparison of ResNet101 on various mammography datasets.

| Dataset | Total Images | Precision | Recall | F1-Score | Accuracy | MCC |
| --- | --- | --- | --- | --- | --- | --- |
| INbreast | 367 | 0.462 | 0.607 | 0.498 | 0.607 | -0.004 |
| CDD-CESM | 1,003 | 0.504 | 0.5 | 0.491 | 0.5 | 0.256 |
| VinDr-Mammo | 19,238 | 0.677 | 0.698 | 0.592 | 0.698 | 0.105 |
| DMID | 390 | 0.55 | 0.25 | 0.234 | 0.25 | 0.155 |
| Mammo-Bench | 34,721 | **0.760** | **0.778** | **0.766** | **0.778** | **0.595** |

### Hierarchical Binary Classification

Next, we performed a two-step binary classification to handle class imbalance. First the model was trained to classify images into normal and abnormal (benign and malignant) cases. Merging benign and malignant classes helped in balancing the classes to a certain extent (Table 3). Next, another model was trained on the correctly predicted abnormal images from the previous step, to classify the images as benign or malignant. It may be noted that the model achieved an accuracy of 0.89 to distinguish between the normal and abnormal cases, and 0.73 to distinguish between Benign and Malignant classes. There is a significant improvement in the recall and precision values of the Benign and Malignant classes compared to 3-class classification results. Notably, performance of the Benign class improved significantly without affecting the performance of other two classes. This again shows the utility of Mammo-Bench in building strategic models with superior performances.

## 5 Discussion

### FAIRness of the Dataset

Mammo-Bench adheres to the FAIR (Findable, Accessible, Interoperable, and Reusable) principles [23]. In terms of Findability, the dataset is described with rich metadata, including detailed information on patient demographics, abnormality types, breast density categories, and BI-RADS classifications. Its structure, with unique image IDs and uniform file structure and paths enhance its findability and organization. Accessibility is ensured through documentation of the dataset’s composition and preprocessing methods, while Interoperability is enhanced by the standardized preprocessing pipeline applied across source datasets. The preprocessing ensures consistent image format and quality. The documentation (including description of data sources, preprocessing methods and annotations) ensures reusability and compatibility across different analysis platforms and machine learning frameworks.

The primary advantage of this dataset lies in its extensive collection of images, which can be utilized for developing and training deep learning models across various detection and classification tasks. It is free of any unlabeled or missing files, ensuring its reliability and value for a wide range of analyses. The dataset’s standardized and user-friendly format further enhances its uniqueness as a comprehensive resource for mammography image analysis. A limitation of this dataset is that class imbalance persists even after combining multiple datasets. This requires the need for data balancing strategies during model training.

## 6 Conclusion

In this paper we present a large-scale, unified mammogram benchmark dataset consisting of 19,731 images. The comprehensive preprocessing pipeline proposed in this work has helped in standardizing the images from diverse sources, and providing regions of interest (ROIs) for the abnormalities in the images and removing irrelevant features. The usability of the proposed dataset is demonstrated by training ResNet101 on it and four other smaller datasets. The organization of the dataset allows for various classification tasks and assesses the impact of breast density and abnormality information on prediction. This makes Mammo-Bench a valuable resource for the development and evaluation of CAD systems. While Mammo-Bench represents a significant step forward in mammography datasets, certain limitations persist, particularly regarding class imbalance and demographic imbalances across different geographical regions ( SupplementaryFigure 1). This may introduce potential biases in the model development and future work may attempt to address these issues. Additionally, a promising direction for future research involves the integration of multimodal data, such as combining mammography images with ultrasound, MRI, or genomic data, to enable personalized and precise screening and propose novel diagnostic strategies.

## Data Availability

All data produced are available online at: https://india-data.org/dataset-details/c86fb00c-0fb8-4e0e-85a2-4d415f9c1ada

## Dataset Availability

Link to the dataset: Mammo-Bench (Login required for access)

## Code Availability

The preprocessing code: https://github.com/Gaurav2543/Mammo-Bench

## Disclaimer

The mammographic images used were sourced from public datasets with appropriate permissions and usage compliance. This dataset is intended for research purposes only and should not be used for direct clinical diagnosis.

## License

Mammo-Bench: A Large-Scale Benchmark Dataset of Mammography Images © 2024 by Gaurav Bhole is licensed under CC BY-NC-SA 4.0. To view a copy of this license, visithttps://creativecommons.org/licenses/by-nc-sa/4.0/

## Author Contributions

G. B. collated and analyzed the data. G.B. was responsible for training the models and writing the code for data handling. S.S. and N.P. supervised the training of the models, and all the technical details. G.B. wrote the initial draft. S.S. and N.P. supported the study, edited, reviewed and worked on the revised manuscript. All authors subsequently critically edited the report. All authors have read and approved the final report.

## Supplementary File

This supplementary material provides additional details about the Mammo-Bench dataset. In Table S1, links for downloading the data from the individual sources are given. The geographical distribution of the data collated in Mammo-Bench is given figure S1. Detailed comparative performance evaluation of ResNet101 on five datasets, namely, Mammo-Bench, INBreast, CDD-CESM, DMID, and VinDr-Mammo is summarized. The information presented here complements the main manuscript by providing details regarding data collection and model performance.

From the class-wise performance across different datasets in Table S2, it may be noted that performance of ResNet101 trained on Mammo-Bench was significantly better when compared to that on other datasets. For normal class, Mammo-Bench achieved high precision (0.865) and recall (0.928), resulting in the best F1-score (0.895), while for malignant class, precision, recall, and F1-score of 0.708, 0.743 and 0.725, respectively, was achieved. Training the model on VinDr-Mammo gave a very good recall (0.99) for normal class, but completely failed to identify malignant class (0.0 for all metrics). Similar poor results are observed on using InBreast and DMID datasets. For benign cases, INbreast showed high recall (0.94), but its overall performance is compromised by poor results in other classes. While DMID showed high precision (0.75) but very low recall (0.12). Mammo-Bench maintained moderate but more balanced performance (precision 0.502, recall 0.369). The overall accuracy of 0.778 and MCC score of 0.595 is significantly higher with Mammo-Bench compared to other datasets. These results indicate that using larger, more diverse training data would lead to a more reliable prediction across all three classes. This balanced performance is crucial for practical clinical applications, where both false positives and false negatives need to be minimized.

**Table S1.** Links to the Individual Datasets.

| Dataset Name | Link |
| --- | --- |
| DDSM | http://www.eng.usf.edu/cvprg/mammography/database.html |
| INbreast | https://www.kaggle.com/datasets/ramanathansp20/inbreast-dataset |
| KAU-BCMD | https://www.kaggle.com/datasets/asmaasaad/king-abdulaziz-university-mammogram-dataset |
| CMMD | https://www.cancerimagingarchive.net/collection/cmmd/ |
| CDD-CESM | https://www.cancerimagingarchive.net/collection/cdd-cesm/ |
| DMID | https://figshare.com/articles/dataset/_b_Digital_mammography_Dataset_for_Breast_Cancer_Diagnosis_Research_DMID_b_DMID_rar/24522883 |

**Figure S1.**
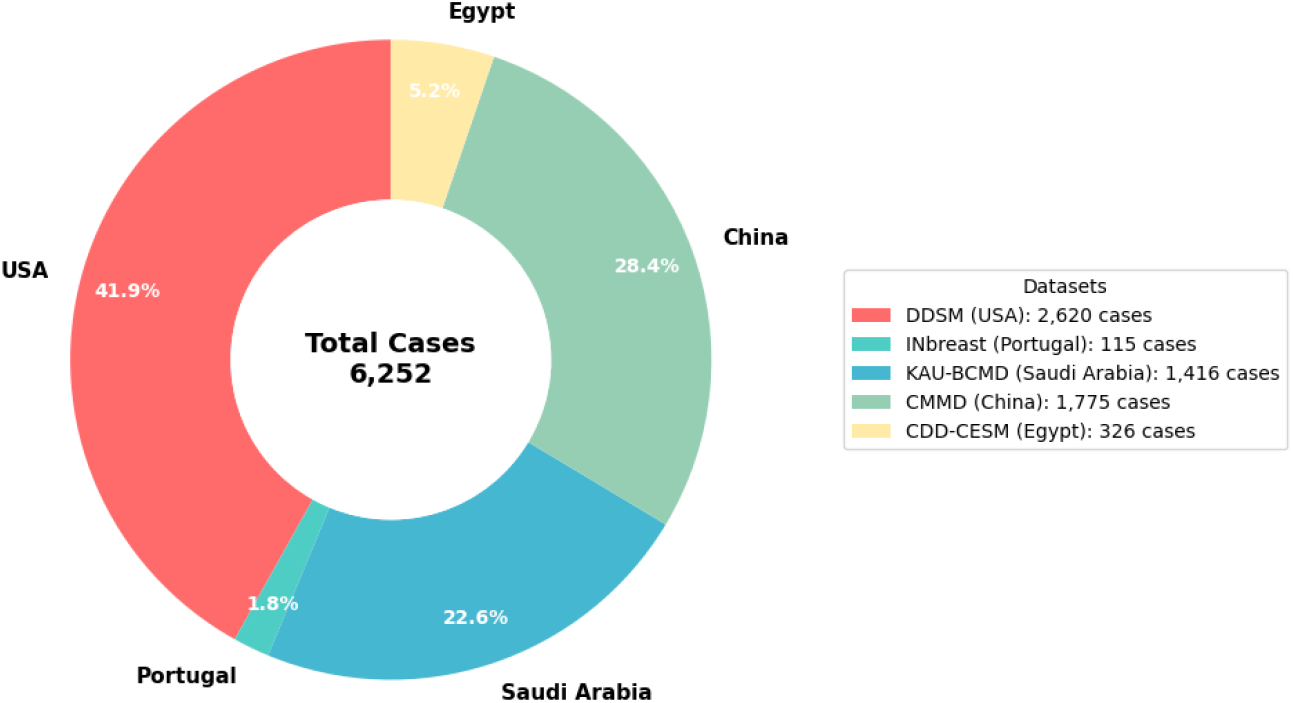
Geographical distribution of the subjects across different datasets included in Mammo-Bench

**Table S2.** Detailed results on a few Individual Datasets.

| Dataset | Class | No. of Images | Precision | Recall | F1-Score | Accuracy | MCC |
| --- | --- | --- | --- | --- | --- | --- | --- |
| INbreast | Normal | 67 | 0.33 | 0.09 | 0.14 | 0.61 | -0.004 |
|  | Benign | 243 | 0.62 | **0.94** | **0.75** |  |  |
|  | Malignant | 57 | 0.00 | 0.00 | 0.00 |  |  |
| CDD-CESM | Normal | 341 | 0.55 | 0.53 | 0.54 | 0.5 | 0.256 |
|  | Benign | 331 | 0.32 | 0.26 | 0.28 |  |  |
|  | Malignant | 331 | 0.57 | 0.69 | 0.62 |  |  |
| DMID | Normal | 212 | 0.63 | 0.18 | 0.28 | 0.25 | 0.155 |
|  | Benign | 154 | **0.75** | 0.12 | 0.21 |  |  |
|  | Malignant | 24 | 0.15 | **1.00** | 0.26 |  |  |
| VinDr-Mammo | Normal | 13,406 | 0.70 | **0.99** | 0.82 | 0.698 | 0.105 |
|  | Benign | 5,606 | 0.63 | 0.04 | 0.08 |  |  |
|  | Malignant | 226 | 0.00 | 0.00 | 0.00 |  |  |
| Mammo-Bench | Normal | 20,646 | **0.865** | 0.928 | **0.895** | **0.778** | **0.595** |
|  | Benign | 6,930 | 0.502 | 0.369 | 0.425 |  |  |
|  | Malignant | 7,145 | **0.708** | 0.743 | **0.725** |  |  |

## Footnotes

* dataset with restricted access

